# Genetic Elucidation of Ultrasonography Fetal Anomalies in Children with Autism Spectrum Disorder

**DOI:** 10.1101/2023.08.05.23293591

**Authors:** Ohad Regev, Apurba Shil, Tal Bronshtein, Amnon Hadar, Gal Meiri, Dikla Zigdon, Analya Michaelovski, Ilan Dinstein, Reli Hershkovitz, Idan Menashe

**Affiliations:** Joyce & Irving Goldman Medical School, Faculty of Health Sciences, Ben-Gurion University of the Negev, Beer-Sheva, Israel; Department of Epidemiology, Biostatistics and Community Health Sciences, Faculty of Health Sciences, Ben-Gurion University of the Negev, Beer-Sheva, Israel; Clalit Health Services, Beer-Sheva, Israel; Division of Obstetrics and Gynecology, Soroka University Medical Center, Beer-Sheva, Israel; Preschool Psychiatric Unit, Soroka University Medical Center, Beer-Sheva, Israel; Child Development Center, Soroka University Medical Center, Beer-Sheva, Israel; Psychology and Brain and Cognition Departments, Ben-Gurion University of the Negev, Beer-Sheva, Israel

## Abstract

Recent evidence suggests that certain fetal anomalies detected upon prenatal ultrasound screenings may be associated with autism spectrum disorder (ASD). In this cross-sectional study, we aimed to identify genetic variants associated with fetal ultrasound anomalies (UFAs) in children with ASD. The study included all children with ASD who are registered in the database of the Azrieli National Center of Autism and Neurodevelopment and for whom both prenatal ultrasound and whole exome sequencing (WES) data were available. We applied our in-house integrative bioinformatics pipeline, *AutScore*, to these WES data to prioritize gene-disrupting variants (GDVs) probably contributing to ASD susceptibily. Univariate statistics and multivariable regression were used to assess the associations between UFAs and GDVs identified in these children. The study sample comprised 126 children, of whom 43 (34.1%) had at least one UFA detected in the prenatal ultrasound scan. A total of 87 candidate ASD genetic variants were detected in 60 children, with 24 (40%) children carrying multiple variants. There was a weak, but significant, correlation between the number of mutations and the number of abnormalities detected in the same children (r = 0.21, *P* = 0.016). Children with UFAs were more likely to have loss-of-function (LoF) mutations (aOR=2.55, 95%CI: 1.13–5.80). This association was particularly noticeable when children with structural anomalies or children with UFAs in their head and brain scans were compared to children without UFAs (any mutation: aOR=8.28, 95%CI: 2.29–30.01; LoF: aOR=5.72, 95%CI: 2.08–15.71 and any mutation: aOR=6.39, 95%CI: 1.34–30.47; LoF: aOR=4.50, 95%CI: 1.32–15.35, respectively). GDVs associated with UFAs were enriched in genes highly expressed across all tissues (aOR=2.76, 95%CI: 1.14–6.68). The results provide valuable insights into the potential genetic basis of prenatal organogenesis abnormalities associated with ASD and shed light on the complex interplay between genetic factors and fetal development.

## INTRODUCTION

Autism spectrum disorder (ASD) is a multifactorial neurodevelopmental disorder with a remarkably heterogeneous clinical presentation and a wide range of associated clinical symptoms ^1–3^. At present, ASD diagnosis is determined after birth ^4^, but a growing body of evidence suggests that in some cases initial signs of ASD can be detected *in utero* ^5–12^. Indeed, several studies have reported that certain fetal abnormalities detected in standard prenatal ultrasound screening are associated with ASD ^5, 7,17, 8–11, 13–16^.

Our group recently completed two studies that explored abnormalities in fetal development associated with ASD. In the first, using prenatal ultrasound biometric data from the second and third trimesters of gestation, we showed that children later diagnosed with ASD had abnormal fetal head growth compared with other fetuses ^5^. In the second study, which focused on prenatal ultrasound data from the late anatomy survey conducted during mid-gestation, we showed that children later diagnosed with ASD had higher rates of fetal structural anomalies and fetal soft markers than their non-ASD counterparts ^11^. This ASD-specific excess of ultrasonography fetal anomalies (UFAs) was mainly seen in the head and brain, the heart, and the urinary system. Taken together, these findings suggest genetic factors or early gestational exposures contributing to the predisposition to abnormal prenatal development of children later diagnosed with ASD.

It is widely accepted that genetic factors play a significant role in ASD susceptibility ^18–22^. In the past two decades, advances in next-generation sequencing (NGS) technologies have facilitated the identification of hundreds of rare genetic variants implicated in ASD susceptibility ^23–27^. Importantly, some of the genetic variants associated with ASD have also been associated with various congenital diseases and malformations. For example, facial and head abnormalities usually occur in ASD-associated genetic syndromes that involve the *CHD8* ^28, 29^ and *ADNP* ^30, 31^ genes and genes in the *22q11.2* region ^32^. Furthermore, comorbidity of ASD and congenital renal abnormalities may be found in people with Phelan-McDermid syndrome ^33, 34^, 17q12 microdeletion syndrome ^13, 17, 35^, or 16q24.2 deletions ^16^, while comorbidity of ASD and congenital heart defects is associated with 22q11.2 deletion syndrome ^32^ and with a set of genes involved in chromatin organization ^36^. Taken together, these findings suggest shared molecular mechanisms underlying both ASD predisposition and abnormal embryonic organogenesis of various body parts.

Despite the emerging literature suggesting abnormal organogenesis and prenatal neurodevelopment in children with ASD and possible genetic etiologies, no studies have investigated the genetic background of prenatal organ development as detected in the fetal ultrasound scans of children later diagnosed with ASD. Herein, we present the first study to examine the association between abnormal fetal development as detected upon fetal ultrasonography and genetic mutations in ASD children.

## MATERIALS AND METHODS

### Study design

We conducted a cross-sectional study comprising all children diagnosed with ASD who are registered in the database of the Azrieli National Center of Autism and Neurodevelopment (ANCAN) ^37^, who are members of Clalit Health Services (CHS; the largest health services provider in Israel), and for whom both prenatal ultrasound and whole exome sequencing (WES) data are available (Figure 1).

**Figure 1.**
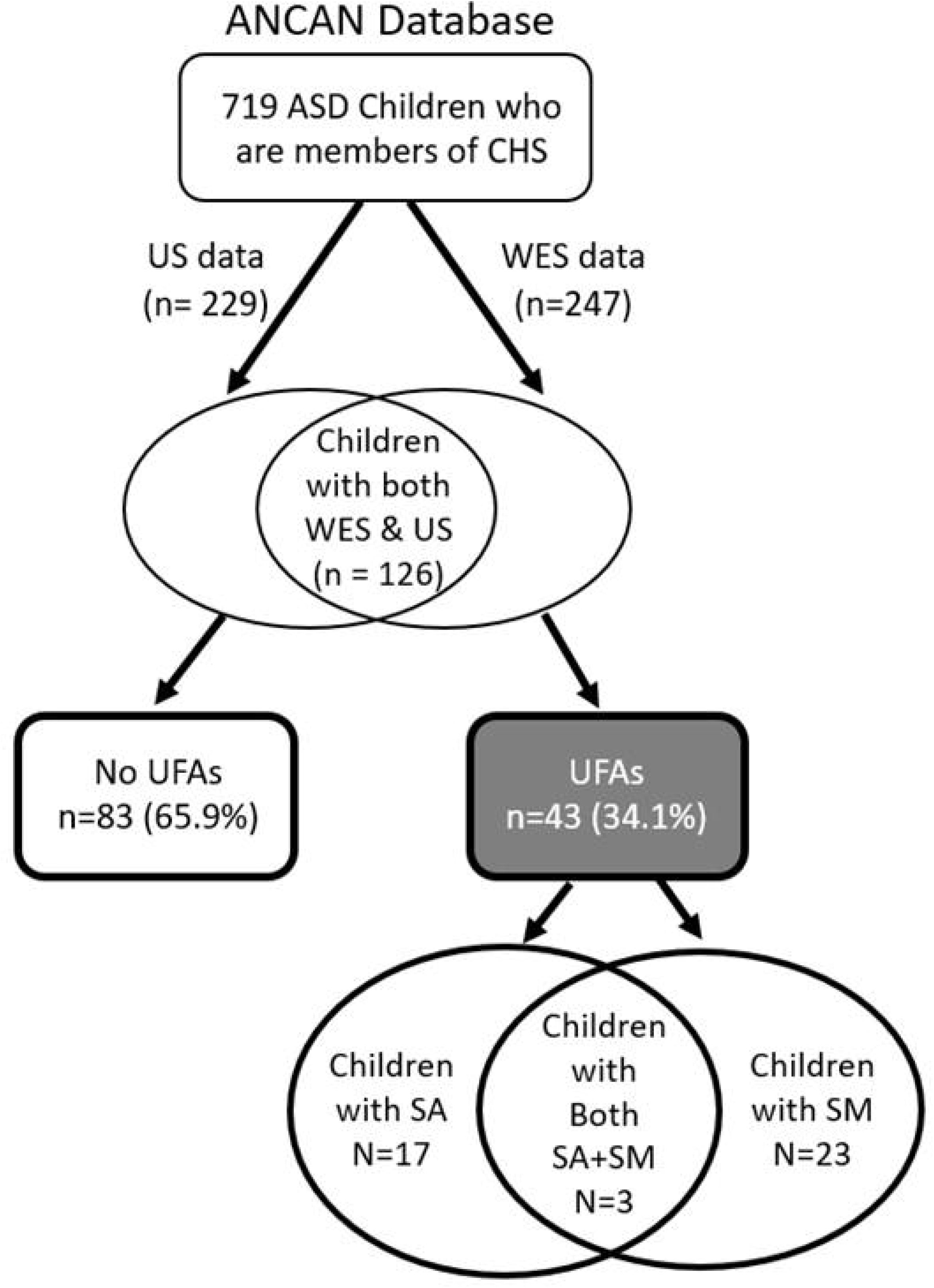
Flowchart of children included in this study. ANCAN: Azrieli National Center for Autism and Neurodevelopment Research; ASD: autism spectrum disorder; CHS: Clalit Health Services; SA: structural anomalies; SM; soft markers; US = ultrasound; WES = whole exome sequencing

### ASD diagnosis

All participants underwent the same diagnosis process, which entailed a comprehensive intake interview (sociodemographic and clinical factors), a behavioral evaluation with ADOS-2 ^38^, and a full neurocognitive assessment using either the Bayley Scales of Infant and Toddler Development - third edition (Bayley-III) ^39^ or the Wechsler Preschool and Primary Scale of Intelligence - version three (WPPSI-III) ^40^, all translated into Hebrew as described previously ^37, 41^. In addition, the level of language development was assessed using the Preschool Language Scale - fifth edition (PLS-5) ^42^. The final diagnosis of ASD was made by a pediatric psychiatrist or neurologist, according to DSM-5 criteria ^43^.

### Fetal ultrasound and birth data

The study is based on prenatal ultrasound data from the fetal anatomy survey that is routinely conducted in Israel during gestational weeks 20–24. The fetal ultrasound data were obtained from all the prenatal ultrasound clinics of the CHS in southern Israel (the intake area for ANCAN). In these clinics, fetal anatomy surveys are performed by experienced physicians, who record fetal anomalies and biometric measures according to standard clinical guidelines ^44, 45^. The ultrasound screening includes the examination of different anatomical landmarks according to the various body systems, including the head, brain, thorax, abdomen, spine, limbs, and umbilical cord. Any abnormalities in each examined organ are classified as structural anomalies or soft markers ^44–46^. Additional measurements of the fetal head and brain include head circumference, biparietal diameter, size of the cisterna magna, and lateral ventricle width ^44, 47^. For the current study, the gestational age of each fetus was calculated from the last menstrual period (LMP) and confirmed by the crown-rump length (CRL) in the ultrasound scan obtained in the first trimester. If the LMP date was not known, gestational age was calculated based on CRL.

UFAs were divided into either soft markers or structural anomalies. The soft markers found in our study included pyelectasis, echogenic intracardiac focus, single umbilical artery, persistent right umbilical vein, echogenic bowel, enlarged fetal stomach, and choroid plexus cyst. The structural anomalies included microcephaly (<3 standard deviations), macrocephaly (>3 standard deviations), ventricular septal defect, ventriculomegaly, mega cisterna magna, single kidney, cystic kidney, dual collection system of the kidneys, and right aortic arch + vascular ring. Birth characteristics of the children included in the study were obtained from the database of the Division of Obstetrics and Gynecology, Soroka University Medical Center (SUMC). The birth characteristics of children born outside of SUMC were obtained from the “Ofek” database, which includes medical data from most hospitals in Israel.

### Identification of candidate ASD genetic variants

WES was performed on DNA purified from saliva samples collected from the affected children and their parents with Genotek OG-500 and OG-575 saliva collection kits, as described previously ^48^. An in-house integrative bioinformatics pipeline, *AutScore* ^49^, was then applied to the WES data to prioritize gene-disrupting variants (GDVs) probably contributing to ASD susceptibily. *AutScore* anlyzes each candidate GDV through a variety of bioinformatics tools and databases and subsequently assigns a summary score to each GDV, based on its predicted clinical pathogenicity, its inheritence characteristics, and the relevance of the affected gene to ASD or other neurodevelopmental disorders. Genetic variants with *AutScore* ≥11 were included in this study as candidate ASD genetic variants.

Storage of the genetic data, data processing, and the detection of GDVs were conducted on a high-performing computer cluster in a Linux environment using Python version 3.5 and R Studio version 1.1.456.

### Statistical analysis

We divided the study sample into children with (Cases) and without (Controls) UFAs in their ultrasonography anatomical surveys and then further divided the UFA group into those with soft markers and those with structural anomalies (Figure 1). Differences in sociodemographic and clinical characteristics between the study groups were tested using appropriate univariate statistics while adjusting for multiple testing using the Bonferroni correction. Spearman’s correlation was used to assess the correlation between the number of mutations and the number of UFAs in the sample. Logistic regression was used to assess the association between the identified GDVs and the existence of any UFA as well as the existence of different types of UFAs while adjusting for the sex of the children.

We used data from the Genotype-Tissue Expression (GTEx) Multi Gene Query in the GTEx Portal ^50^ to assess the level of transcripts per million (TPM) of genes with identified variants in different tissues. Then, we applied the GTEx Multi Gene Query genetic clustering ^50, 51^ to identify four clusters of gene expression patterns. Finally, we assessed the association of different types of UFAs with these clusters using logistic regression, as described above.

All statistical analyses were conducted using SPSS Statistics V. 25 and R software. A two-sided test significance level of 0.05 was used throughout the entire study.

## RESULTS

### Sample characteristics

The study sample included 126 children with ASD (6 sibling pairs and 114 singletons) for whom both WES and ultrasound data were available (Figure 1). The children in the study sample had a relatively lower age at diagnosis and a higher ADOS score compared to other children with ASD in the ANCAN database (Supplementary Table S1). Of the 126 children, 43 (34.1%) had at least one reported UFA in their anatomical ultrasonography surveys; of these 43 children, in turn, 17 (13.5%) had UFAs defined as structural anomalies, 23 (18.3%) had UFAs defined as soft markers, and 3 children (2.4%) had both structural anomalies and soft markers. In the remaining 83 children (65.9%), no UFAs were detected in the anatomical ultrasonography survey.

Table 1 presents the sociodemographic and clinical characteristics of the study sample, including a comparison of these characteristics between children with and without UFAs. The study sample comprised 77.8% males and 73.8% children of Jewish ethnicity, without significant differences between cases and controls. There were also no significant differences between children with and without UFAs in terms pregnancy, birth, and ultrasound details, and clinical severity characteristics. Of note, children in the soft marker group had slightly higher, although not statistically significant, cognitive and language abilities compared to children with structural markers and children without UFAs (82.4±19.1 vs. 74.8±13.3 and 74.8±16.4, respectively, for IQ score, and 82.2±25.3 vs. 57.7±11.5 and 61.7±20.6, respectively, for PLS score).

**Table 1.**
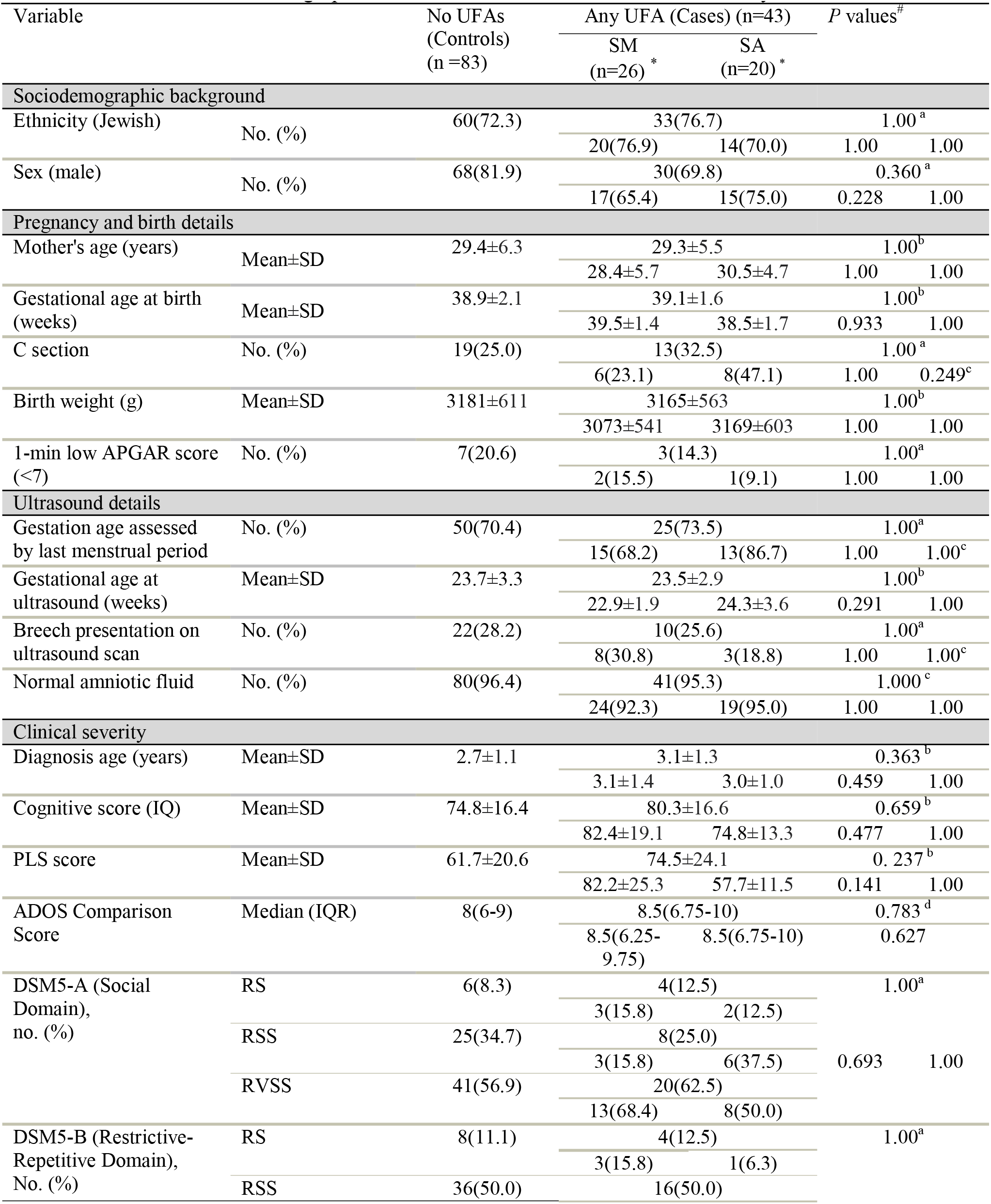
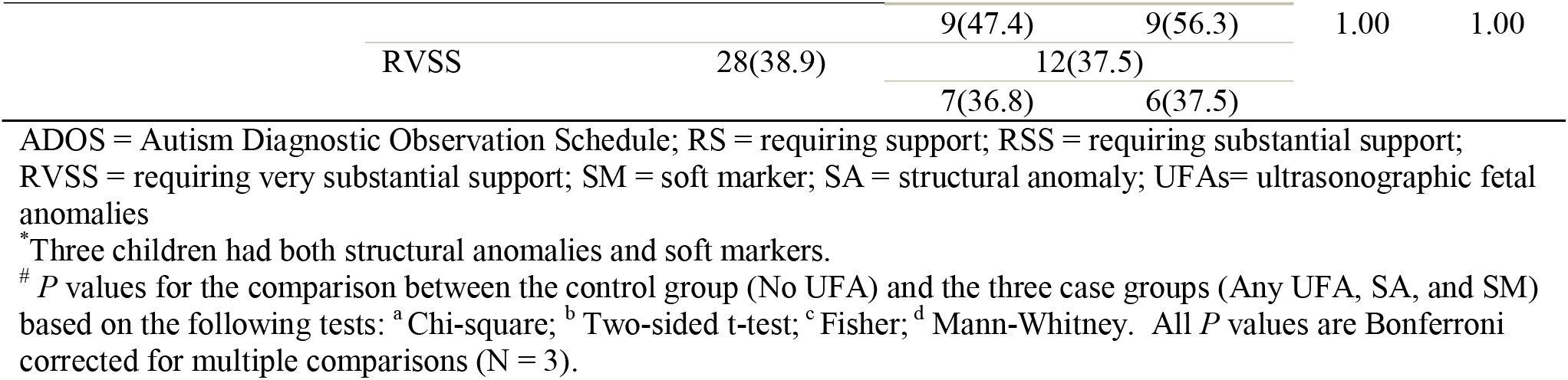
Clinical and sociodemographic characteristics for children included in study cohort.

### Genetic findings

Overall, 87 GDVs with *AutScore* ≥11 were identified in 60 children in the study sample (children with both ultrasound and WES data). Of these, 23 children (38.3%) carried more than one GDV. A list of these GDVs, their genetic characteristics, and their associated UFAs is given in Supplementary Table S2. Seventy-two of the detected GDVs were inherited (69 dominant, and 3 recessive), while 15 GDVs were de-novo variants. In terms of functional consequences, 43 variants were classified as loss-of-function (LoF) mutations (i.e., frameshift, stop gain/loss, and splice acceptor/donor variants), while the remaining 44 variants were missense mutations. The genetic characteristics of GDVs with *AutScore* ≥11 did not vary significantly between children with and without ultrasound data (Supplementary Table S3).

### Association between UFAs and candidate ASD genetic variants

UFAs in children with ASD were associated with different characteristics of detected GDVs (Table 2). Specifically, children with UFAs were 2.5 times more likely to carry LoF mutations than children without UFAs (aOR = 2.55, 95%CI: 1.13–5.80). This association was particularly noticeable when comparing children with structural anomalies or children with head and brain UFAs to children without UFAs (any mutation: aOR = 8.28, 95%CI: 2.29-30.01; LoF: aOR = 5.72, 95%CI: 2.08–15.71 and any mutation: aOR = 6.39, 95%CI: 1.34–30.47; LoF: aOR = 4.50, 95%CI: 1.32–15.35, respectively). Of note, given the significant overlap between structural anomalies and head and brain UFAs (Kappa = 0.38, *P* value <0.001), the significant associations of genetic variants with these types of UFA are not mutually exclusive. In addition, there was a weak, but statistically significant, correlation between the number of detected GDVs and the number of detected UFAs in each child (r = 0.21, *P* = 0.016).

**Table 2.**
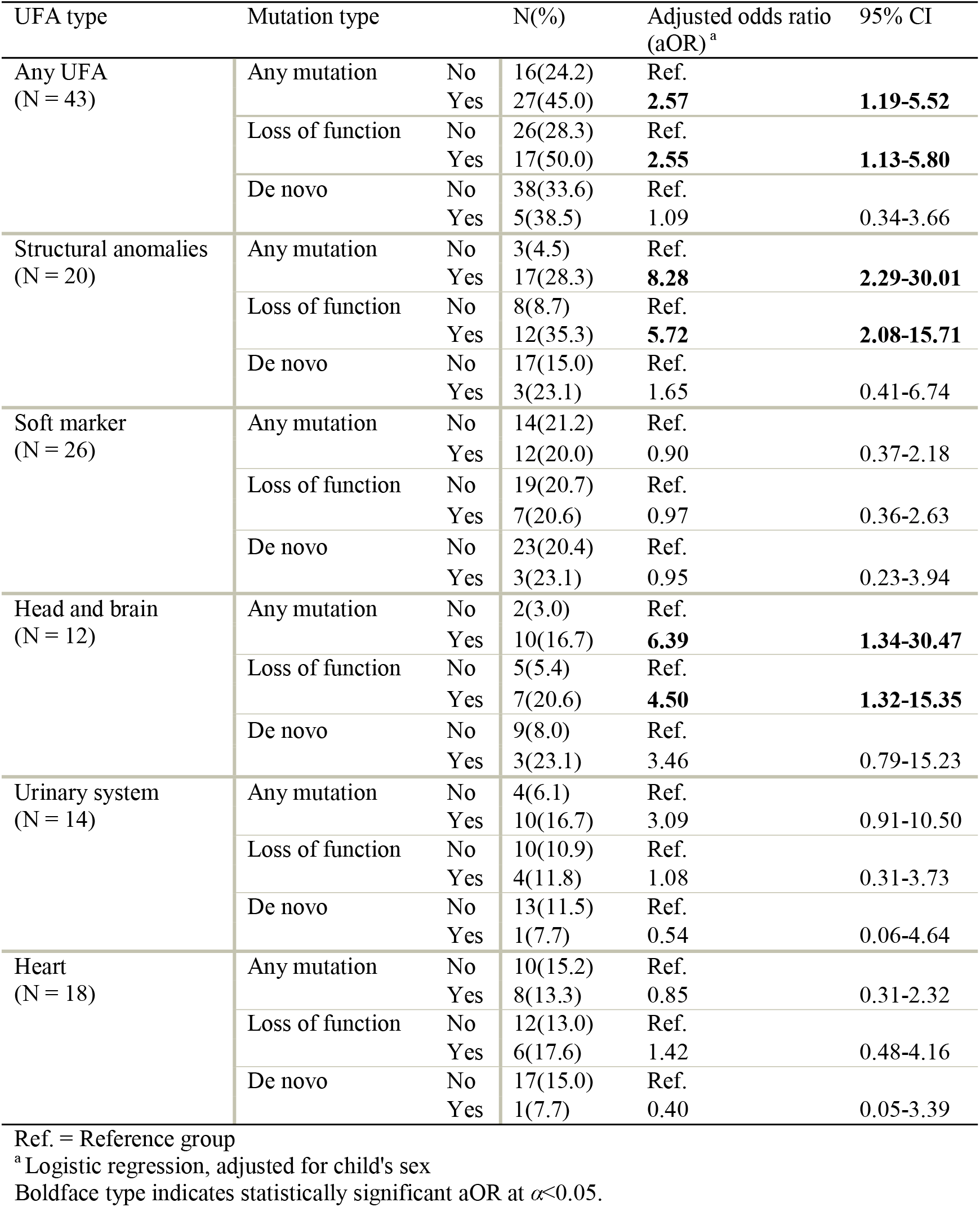
Association between UFAs and genetic mutations.

| **Table 2.** Association between UFAs and genetic mutations |  |  |  |  |  |
| --- | --- | --- | --- | --- | --- |
| UFA type | Mutation type |  | N(%) | Adjusted odds ratio (aOR)<sup>a</sup> | 95% CI |
| Any UFA<br>(N = 43) | Any mutation | No | 16(24.2) | Ref. |  |
|  |  | Yes | 27(45.0) | **2.57** | **1.19-5.52** |
|  | Loss of function | No | 26(28.3) | Ref. |  |
|  |  | Yes | 17(50.0) | **2.55** | **1.13-5.80** |
|  | De novo | No | 38(33.6) | Ref. |  |
|  |  | Yes | 5(38.5) | 1.09 | 0.34-3.66 |
| Structural anomalies<br>(N = 20) | Any mutation | No | 3(4.5) | Ref. |  |
|  |  | Yes | 17(28.3) | **8.28** | **2.29-30.01** |
|  | Loss of function | No | 8(8.7) | Ref. |  |
|  |  | Yes | 12(35.3) | **5.72** | **2.08-15.71** |
|  | De novo | No | 17(15.0) | Ref. |  |
|  |  | Yes | 3(23.1) | 1.65 | 0.41-6.74 |
| Soft marker<br>(N = 26) | Any mutation | No | 14(21.2) | Ref. |  |
|  |  | Yes | 12(20.0) | 0.90 | 0.37-2.18 |
|  | Loss of function | No | 19(20.7) | Ref. |  |
|  |  | Yes | 7(20.6) | 0.97 | 0.36-2.63 |
|  | De novo | No | 23(20.4) | Ref. |  |
|  |  | Yes | 3(23.1) | 0.95 | 0.23-3.94 |
| Head and brain<br>(N = 12) | Any mutation | No | 2(3.0) | Ref. |  |
|  |  | Yes | 10(16.7) | **6.39** | **1.34-30.47** |
|  | Loss of function | No | 5(5.4) | Ref. |  |
|  |  | Yes | 7(20.6) | **4.50** | **1.32-15.35** |
|  | De novo | No | 9(8.0) | Ref. |  |
|  |  | Yes | 3(23.1) | 3.46 | 0.79-15.23 |
| Urinary system<br>(N = 14) | Any mutation | No | 4(6.1) | Ref. |  |
|  |  | Yes | 10(16.7) | 3.09 | 0.91-10.50 |
|  | Loss of function | No | 10(10.9) | Ref. |  |
|  |  | Yes | 4(11.8) | 1.08 | 0.31-3.73 |
|  | De novo | No | 13(11.5) | Ref. |  |
|  |  | Yes | 1(7.7) | 0.54 | 0.06-4.64 |
| Heart<br>(N = 18) | Any mutation | No | 10(15.2) | Ref. |  |
|  |  | Yes | 8(13.3) | 0.85 | 0.31-2.32 |
|  | Loss of function | No | 12(13.0) | Ref. |  |
|  |  | Yes | 6(17.6) | 1.42 | 0.48-4.16 |
|  | De novo | No | 17(15.0) | Ref. |  |
|  |  | Yes | 1(7.7) | 0.40 | 0.05-3.39 |

Ref. = Reference group
<sup>a</sup>Logistic regression, adjusted for child's sex
Boldface type indicates statistically significant aOR at $\alpha < 0.05$ .

### Association between UFAs and gene expression patterns

Figure 2 shows the tissue expression patterns of genes affected by the detected GDVs according to the GTEx Portal ^50^. We selected the largest four clusters of co-expressed genes (≥10 genes per cluster) and examined their association with the detected UFAs in the study sample (Table 3). The genes of *cluster 4* that are highly expressed across all tissues were significantly more prevalent in children with any UFA (aOR=2.76, 95%CI: 1.14–6.68), and specifically in children with structural anomalies (aOR = 4.07, 95%CI: 1.47–11.30) and UFAs in the head and brain (aOR = 4.50, 95%CI: 1.31–15.42). Genes in *cluster 3*, which were characterized by moderate expression across all tissues, were more likely to be mutated in children with UFAs, specifically soft markers, in the urinary system, but these associations were not statistically significant (aOR = 2.73 95%CI: 0.86–8.62; and aOR = 3.31, 95%CI: 0.89–12.38, respectively).

**Figure 2.**
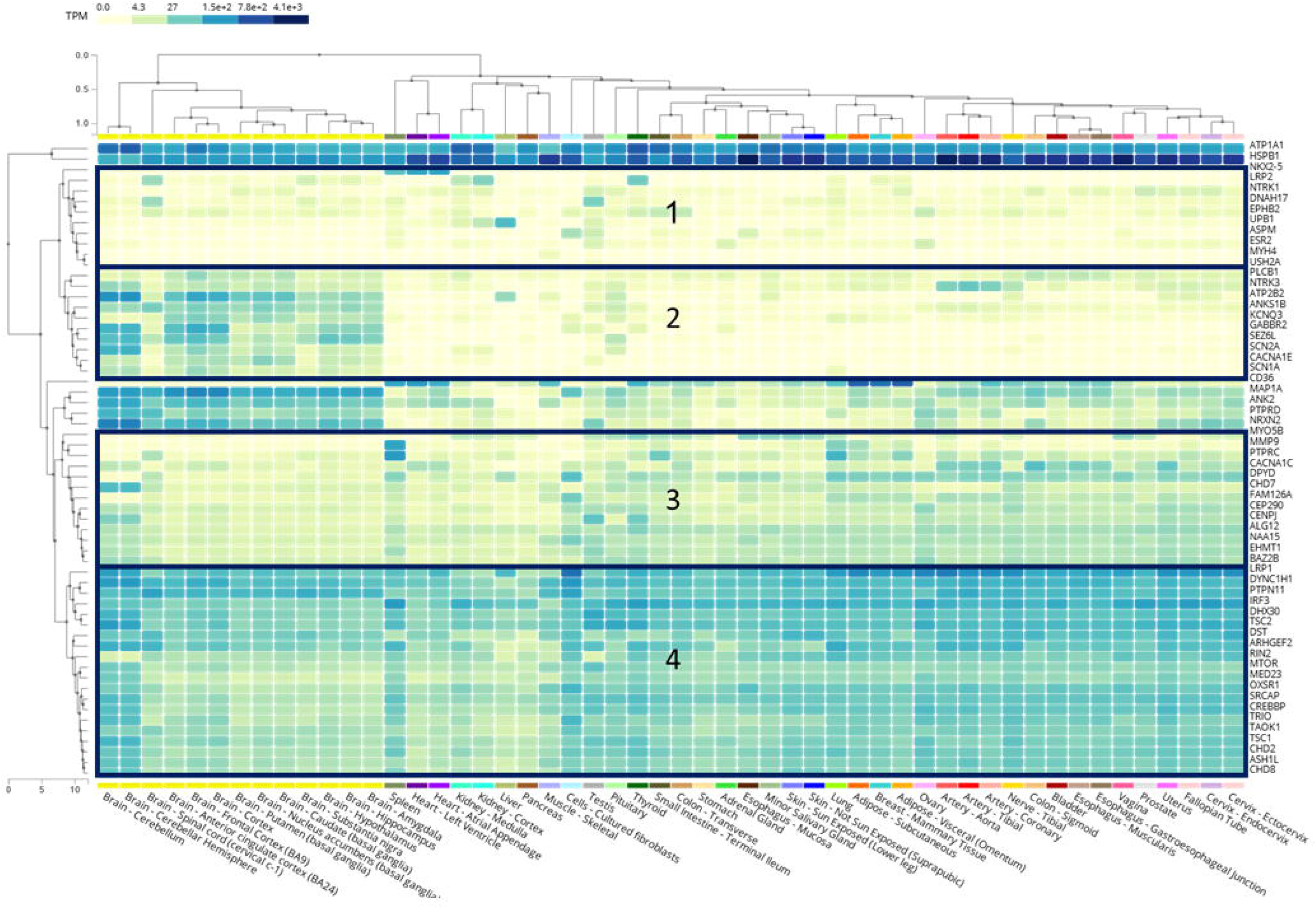
Heatmap of gene expression in different organs. The numbers represent the 4 gene clusters.

**Table 3.**
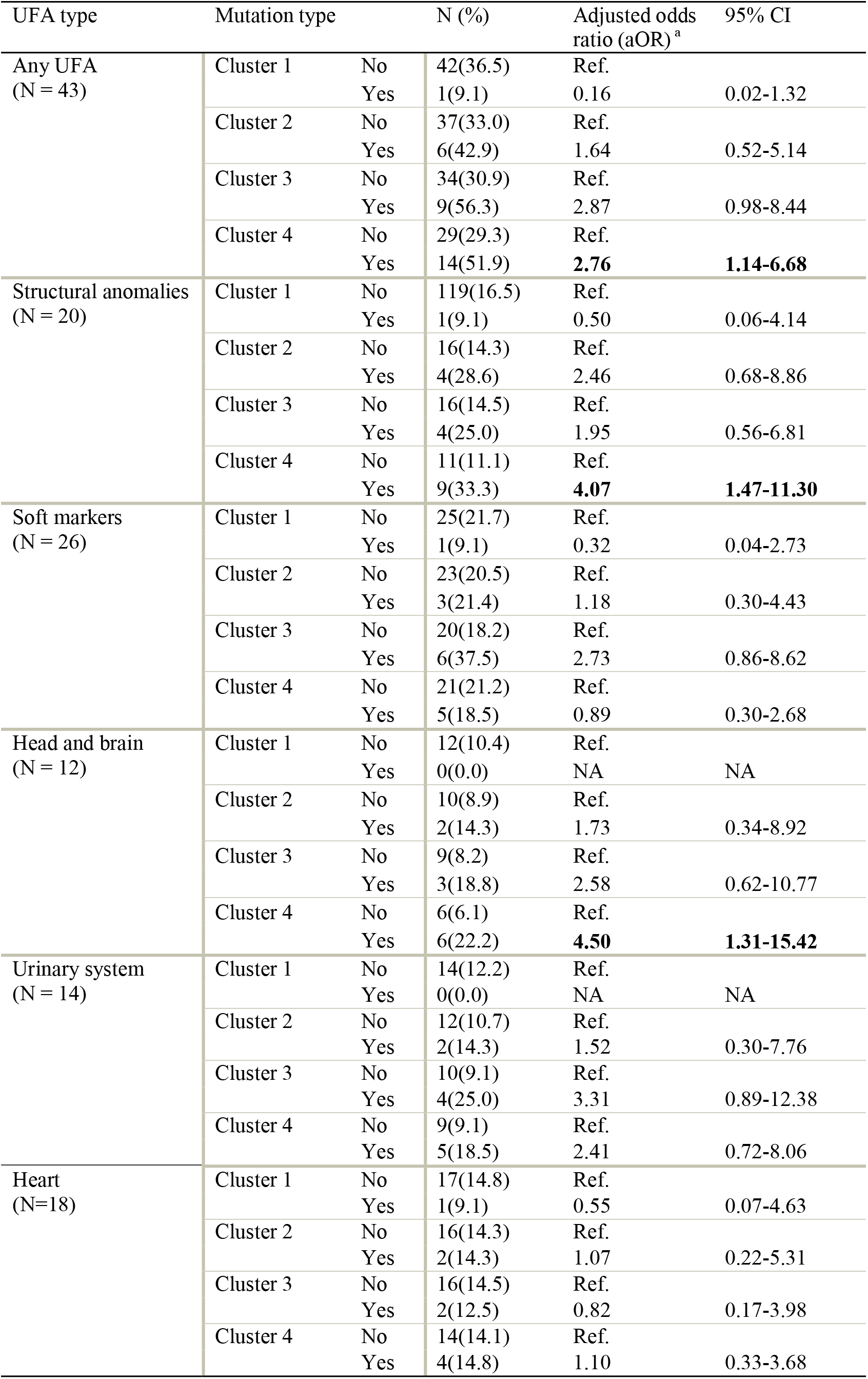
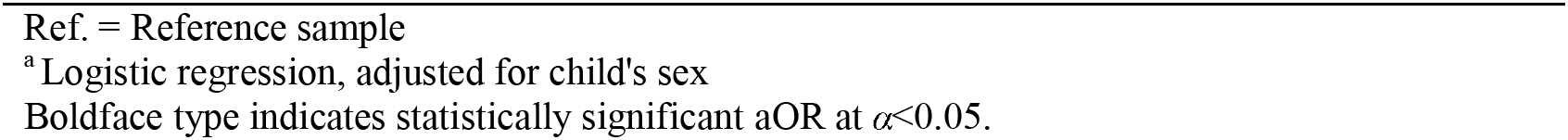
Association between UFAs and genetic clusters.

| **Table 3.** Association between UFAs and genetic clusters |  |  |  |  |  |
| --- | --- | --- | --- | --- | --- |
| UFA type | Mutation type |  | N (%) | Adjusted odds ratio (aOR)<sup>a</sup> | 95% CI |
| Any UFA<br>(N = 43) | Cluster 1 | No | 42(36.5) | Ref. |  |
|  |  | Yes | 1(9.1) | 0.16 | 0.02-1.32 |
|  | Cluster 2 | No | 37(33.0) | Ref. |  |
|  |  | Yes | 6(42.9) | 1.64 | 0.52-5.14 |
|  | Cluster 3 | No | 34(30.9) | Ref. |  |
|  |  | Yes | 9(56.3) | 2.87 | 0.98-8.44 |
|  | Cluster 4 | No | 29(29.3) | Ref. |  |
|  |  | Yes | 14(51.9) | **2.76** | **1.14-6.68** |
| Structural anomalies<br>(N = 20) | Cluster 1 | No | 119(16.5) | Ref. |  |
|  |  | Yes | 1(9.1) | 0.50 | 0.06-4.14 |
|  | Cluster 2 | No | 16(14.3) | Ref. |  |
|  |  | Yes | 4(28.6) | 2.46 | 0.68-8.86 |
|  | Cluster 3 | No | 16(14.5) | Ref. |  |
|  |  | Yes | 4(25.0) | 1.95 | 0.56-6.81 |
|  | Cluster 4 | No | 11(11.1) | Ref. |  |
|  |  | Yes | 9(33.3) | **4.07** | **1.47-11.30** |
| Soft markers<br>(N = 26) | Cluster 1 | No | 25(21.7) | Ref. |  |
|  |  | Yes | 1(9.1) | 0.32 | 0.04-2.73 |
|  | Cluster 2 | No | 23(20.5) | Ref. |  |
|  |  | Yes | 3(21.4) | 1.18 | 0.30-4.43 |
|  | Cluster 3 | No | 20(18.2) | Ref. |  |
|  |  | Yes | 6(37.5) | 2.73 | 0.86-8.62 |
|  | Cluster 4 | No | 21(21.2) | Ref. |  |
|  |  | Yes | 5(18.5) | 0.89 | 0.30-2.68 |
| Head and brain<br>(N = 12) | Cluster 1 | No | 12(10.4) | Ref. |  |
|  |  | Yes | 0(0.0) | NA | NA |
|  | Cluster 2 | No | 10(8.9) | Ref. |  |
|  |  | Yes | 2(14.3) | 1.73 | 0.34-8.92 |
|  | Cluster 3 | No | 9(8.2) | Ref. |  |
|  |  | Yes | 3(18.8) | 2.58 | 0.62-10.77 |
|  | Cluster 4 | No | 6(6.1) | Ref. |  |
|  |  | Yes | 6(22.2) | **4.50** | **1.31-15.42** |
| Urinary system<br>(N = 14) | Cluster 1 | No | 14(12.2) | Ref. |  |
|  |  | Yes | 0(0.0) | NA | NA |
|  | Cluster 2 | No | 12(10.7) | Ref. |  |
|  |  | Yes | 2(14.3) | 1.52 | 0.30-7.76 |
|  | Cluster 3 | No | 10(9.1) | Ref. |  |
|  |  | Yes | 4(25.0) | 3.31 | 0.89-12.38 |
|  | Cluster 4 | No | 9(9.1) | Ref. |  |
|  |  | Yes | 5(18.5) | 2.41 | 0.72-8.06 |
| Heart<br>(N=18) | Cluster 1 | No | 17(14.8) | Ref. |  |
|  |  | Yes | 1(9.1) | 0.55 | 0.07-4.63 |
|  | Cluster 2 | No | 16(14.3) | Ref. |  |
|  |  | Yes | 2(14.3) | 1.07 | 0.22-5.31 |
|  | Cluster 3 | No | 16(14.5) | Ref. |  |
|  |  | Yes | 2(12.5) | 0.82 | 0.17-3.98 |
|  | Cluster 4 | No | 14(14.1) | Ref. |  |
|  |  | Yes | 4(14.8) | 1.10 | 0.33-3.68 |

Ref. = Reference sample
<sup>a</sup> Logistic regression, adjusted for child's sex
Boldface type indicates statistically significant aOR at $\alpha < 0.05$ .

## Discussion

In this study, we examined the association between UFAs and GDVs in children with ASD. We found that UFAs were detected during the standard fetal anatomical survey in approximately one-third of children with ASD and, particularly, that structural anomalies in the head and brain were associated with mutations in candidate ASD genes. These findings are consistent with the results of multiple genetic studies reporting a probable genetic cause of different head and brain congenital anomalies in ASD children. For example, three children with fetal macrocephaly (a structural anomaly) in our study sample had genetic mutations in genes previously linked to this phenotype. One of these genes is *CEP290,* a gene linked to Joubert Syndrome, which has been associated with multiple fetal anomalies, including macrocephaly, increased biparietal diameter, and ventriculomegaly, on fetal MRI and ultrasound ^52^. In addition, two children in our sample, one with a mutation in *PTPN11*, and the other with mutations in *TSC2* and SRCAP, also exhibited macrocephaly (a structural anomaly), as reported previously ^53–56^. Furthermore, one child in our sample with a de-novo mutation in *MTOR,* which is a key member in the mTOR pathway that regulates cell growth and proliferation ^57, 58^, also exhibited fetal microcephaly (a structural anomaly). It has been reported for animal models that alterations in the mTOR pathway lead to microcephaly during prenatal life as a result of reduced cell number and cell size^57^. Moreover, a recent study on ASD children found an increased risk of microcephaly in mTOR variant carriers as compared to carriers of mutations in non-mTOR genes ^58^. Finally, three children with ventriculomegaly (a structural anomaly) in our sample had genetic mutations in *DPYD, NAA15*, and *SEZL6*. Consistent with these findings, ventriculomegaly has previously been reported in a child with a *DPYD* mutation ^59^ as well as in children with a 4q31.1 deletion and 16p11.2 duplications, which include the *NAA15* and *SEZL6* genes, respectively ^60, 61^.

Our sample included children from several multiplex ASD families. For two siblings with ASD with the same missense/dominant genetic variant in *ANK2*, there were UFAs in their urinary systems. The scan of the older child, who also had a LoF mutation in *MMP9*, exhibited pyelectasis (a soft marker), while that of the younger one showed a polycystic kidney (a structural anomaly). *ANK2* mutations are associated with an increased number of excitatory synapses with augmented axonal branching, which supports the presence of altered connectivity and penetrant behavioral impairments in humans carrying the ASD-related *ANK2* mutation ^62^. In addition, *ANK2* is located on the 4q deletion, previously linked to congenital anomalies affecting different body systems, including developmental delay, cardiac involvement and polycystic kidney or isolated kidney ^63^. Importantly, *MMP9,* which was disrupted in one of these siblings, is known to be involved in prenatal kidney organogenesis and thus may contribute to the upper urinary tract dilatation ^64, 65^. In another multiplex family, the fetal ultrasound scan for a child with missense/dominant mutations in both *CHD7* and *CHD8* genes showed a single kidney (a structural anomaly). For this child, two older siblings with fetal ultrasonography hydronephrosis and dilated bowels died shortly after birth. *CHD8* and its paralog *CHD7* have been previously linked to the CHARGE syndrome in which renal anomalies have been described ^12, 66^. In addition, in studies on both animals and humans, mutations in *CHD8* were characterized by macrocephaly due to expansion of the forebrain/midbrain and a number of distinct facial characteristics ^28^, as well as genitourinary abnormalities^67^.

We showed that the association between UFAs and GDVs is particularly relevant to genes that are broadly expressed in all body tissues (*cluster 4* in our analysis). Previous studies indicated that most early prenatal ASD regulatory genes are broadly expressed in multiple organs in addition to the developing prenatal brain, suggesting that broadly expressed regulatory genes might be a major driver of prenatal neural maldevelopment in ASD and might also affect the development and function of other organs and tissues ^6, 68–70^. Fifteen of the twenty genes comprising *cluster 4* in our analysis (75%) were also reported in ASD genetic studies to be broadly expressed during prenatal life in the fetal body and the fetal brain, causing disruptions of cell proliferation, neurogenesis, migration, and cell fate, and consequently affecting prenatal brain development ^6, 70^ These findings suggest that mutations affecting normal organ development are predominantly expressed during prenatal life and lead to abnormal neurodevelopment and organogenesis, which can be detected on prenatal ultrasound.

We also noticed a weak, but statistically significant, correlation between the number of UFAs and number of GDVs in our sample. Such a correlation may be indicative of a cumulative effect of genetic variations on fetal development. Multiple genetic mutations may lead to more pronounced disruptions of organogenesis, resulting in multiple UFAs. This conclusion highlights the complexity of genetic factors involved in prenatal organ development and suggests that the risk of UFAs in ASD may be influenced by a combination of genetic factors rather than individual mutations alone. Indeed, previous studies found that ASD children with multiple mutations in known ASD genes demonstrate more severe ASD phenotypes (e.g., worse cognitive ability, symptom impairment, and more seizures), suggesting that multiple gene-disruptive events may co-occur in probands and act synergistically or additively to lead to a more severe phenotype ^56, 71, 72^. In addition, a study that explored prenatal-stage regulatory ASD risk genes showed that many of them are upstream regulators of key signaling pathways (e.g., RAS/ERK, PI3K/AKT, WNT/β-catenin) and that the degree of dysregulation in these signaling pathways is associated with the severity of ASD symptoms^6, 73^. Thus, higher dysregulation in the signaling pathways due to multiple genetic mutations may cause more severe ASD alongside higher rates of abnormal organ development.

## Limitations

This study has three main limitations. First, most (∼80%) of the GDVs included in our analyses were inherited from one parent (dominant inheritance). Although the contribution of heritable factors to ASD susceptibility is well established ^74, 75^, evidence for specific dominantly inherited mutations that are robustly associated with ASD is limited. Nonetheless, we decided to include inherited dominant mutations in our analyses with the aim to increase the number of GDVs and subsequently the statistical power of this study. Of note, the associations found between dominantly inherited variants and UFAs in this study were also seen for de-novo variants and UFAs, but the latter did not reach statistical significance due to their small numbers. Although there is some evidence of the role of inherited dominant mutations in ASD susceptibility, further studies are needed to confirm the clinical relevance of these variants to ASD. Second, the study relied on a retrospective analysis of prenatal ultrasound data, which may introduce inherent biases and limitations in the accuracy of UFA detection. Prospective studies with standardized protocols for ultrasound assessment would provide more reliable data on prenatal organ development in ASD. Finally, our study is focused on a sample of the Israeli population, which may limit the generalizability of the findings to other populations.

## Conclusions

Our study suggests that there are specific genetic variations that probably underlie abnormal fetal development in children with ASD. Thus, it provides valuable insights into the potential genetic basis of prenatal organogenesis abnormalities associated with ASD and sheds light on the complex interplay between genetic factors and fetal development.

## Data Availability

All data produced in the present study are available upon reasonable request to the authors

## Acknowledgments

This study was supported by a grant from the Israel Science Foundation (1092/21).

The study was approved by the SUMC Ethics Committee per the Declaration of Helsinki SOR 295-18.

We thank Mrs. Inez Mureinik for critical reviewing and editing of the manuscript.

This study was conducted in partial fulfillment of the requirements for the MD, PhD degree in medicine awarded by the Joyce & Irving Goldman Medical School, Faculty of Health Sciences, Ben-Gurion University of the Negev.

The article has been previously posted in MedRxiv preprint server.

