## Supplementary Materials for "Genetic Elucidation of Ultrasonography Fetal Anomalies in Children with Autism Spectrum Disorder"

| **Supplementary Table S1.** Clinical and sociodemographic characteristics of ASD children | | | |
| --- | --- | --- | --- |
| *P* value | Excluded  (n = 593) | Study cohort  (n = 126) | Variable |
| 0.279 ^a^ | 467(78.2) | 93(73.8) | Jewish, no. (%) |
| 0.317 ^a^ | 472(78.7) | 94(74.6) | Male, no. (%) |
| 0.090 ^a^ | 152(33.9) | 32(29.1) | Special education setting, no. (%) |
| 0.387 ^b^ | 31.0±6.1 | 30.4±6.0 | Maternal age, mean±SD, years |
| 0.241 ^b^ | 34.3±7.6 | 33.4±7.7 | Paternal age, mean±SD, years |
| **<0.001** **^b^** | 3.3±1.5 | 2.8±1.3 | Diagnosis age, mean±SD, years |
| 0.703 ^b^ | 76.1±16.6 | 77.0±16.4 | Cognitive score (IQ), mean±SD |
| **0.023 ^c^** | 7(6-9) | 8(6-9) | ADOS Comparison Score, median (IQR) |
| ^a^ Chi-square; ^b^ Two-sided t-test; ^c^ Mann-Whitney U test  Boldface type indicates statistically significant aOR at α<0.05. | | | |

| **Supplementary Table S3.** Genetic characteristics of ASD children | | | |
| --- | --- | --- | --- |
| *P* value | Children without  ultrasound data  (n = 121) ^a^ | Children with ultrasound data  (n = 126)^a^ | Variable |
| 0.833 ^b^ | 56(46.3) | 60(47.6) | Any mutation |
| 0.414 ^b^ | 41(33.9) | 49(38.9) | Dominant |
| 0.917 ^b^ | 12(9.9) | 13(10.3) | De novo |
| 0.175 ^b^ | 7(5.8) | 3(2.4) | Recessive |
| 0.490 ^c^ | 1(0.8) | 0(0.0) | X-linked |
| 0.814 ^b^ | 30(2.9) | 34(27.2) | LoF |
| ^a^  Values are no. (%);  ^b^ Chi-square; ^c^ Fisher exact test | | | |
